# Interpretable gender classification from retinal fundus images using BagNets

**DOI:** 10.1101/2021.06.21.21259243

**Authors:** Indu Ilanchezian, Dmitry Kobak, Hanna Faber, Focke Ziemssen, Philipp Berens, Murat Seçkin Ayhan

## Abstract

Deep neural networks (DNNs) are able to predict a person’s gender from retinal fundus images with high accuracy, even though this task is usually considered hardly possible by ophthalmologists. Therefore, it has been an open question which features allow reliable discrimination between male and female fundus images. To study this question, we used a particular DNN architecture called BagNet, which extracts local features from small image patches and then averages the class evidence across all patches. The BagNet performed on par with the more sophisticated Inception-v3 model, showing that the gender information can be read out from local features alone. BagNets also naturally provide saliency maps, which we used to highlight the most informative patches in fundus images. We found that most evidence was provided by patches from the optic disc and the macula, with patches from the optic disc providing mostly male and patches from the macula providing mostly female evidence. Although further research is needed to clarify the exact nature of this evidence, our results suggest that there are localized structural differences in fundus images between genders. Overall, we believe that BagNets may provide a compelling alternative to the standard DNN architectures also in other medical image analysis tasks, as they do not require post-hoc explainability methods.

## 1 Introduction

In recent years, deep neural networks (DNNs) have achieved physician-level accuracy in various image-based medical tasks, e.g. in radiology [21], dermatology [10], pathology [15] and ophthalmology [12, 7]. Moreover, in some cases DNNs have been shown to have good performance in tasks that are not straightforward for physicians: for example, they can accurately predict the gender from retinal images [25]. As this task is typically not clinically relevant, ophthalmologists are not explicitly trained for it. Nevertheless, the comparably poor performance of ophthalmologists at this task suggests that gender differences in fundus images are not obvious or salient. Even though saliency maps used by [25] and follow-up studies [9, 5] have tentatively pointed at the optic disc, the macula, and retinal blood vessels as candidate regions for gender-related anatomical differences in fundus images, conclusive evidence is still lacking. Therefore the high gender prediction performance of DNNs has created lots of interest in the medical imaging community as one hope for DNNs is to unravel biomarkers that are not easily found by humans. Here, we performed a proof of principle study to make progress on the question of how DNNs are able to detect gender differences in retinal fundus. Our contribution is twofold: we (1) introduced BagNets [3] — a ‘local’ variant of the ResNet50 architecture [13] — as an interpretable-by-design architecture for image analysis in ophthalmology and (2) used them to narrow down the hypothesis space for question at hand.

We trained the BagNets on a large collection of retinal fundus images obtained from the UK Biobank [27] (Fig. 1a). BagNets use a linear classifier on features extracted from image patches to compute local evidence for each class, which is then averaged over space to form the final prediction, without considering any global relationships. Thus, BagNets resemble ‘bag-of-features’ models popular before deep learning [23]. Despite this simple bag-of-features approach, the BagNet performed on par with an Inception-v3 network in terms of gender prediction accuracy, indicating that gender can be determined from the local characteristics of the fundus image. Also, the BagNet architecture naturally allowed to construct saliency maps to highlight the most informative regions for gender prediction in the retina (Fig. 1b). We found that the macula contained most distinctive female patches, while the optic disk contained male ones. In addition, we showed that the decision of the BagNet was not simply caused by some exclusively female or male patches in the images, but rather by a change in both frequency and the degree of ‘femaleness’ or ‘maleness’ of individual patches. Overall, we argue that BagNets can be useful in medical imaging applications including both disease diagnosis and biomarker discovery, thanks to interpretability provided by their local architecture. Our code will be available upon publication.

**Fig. 1:**
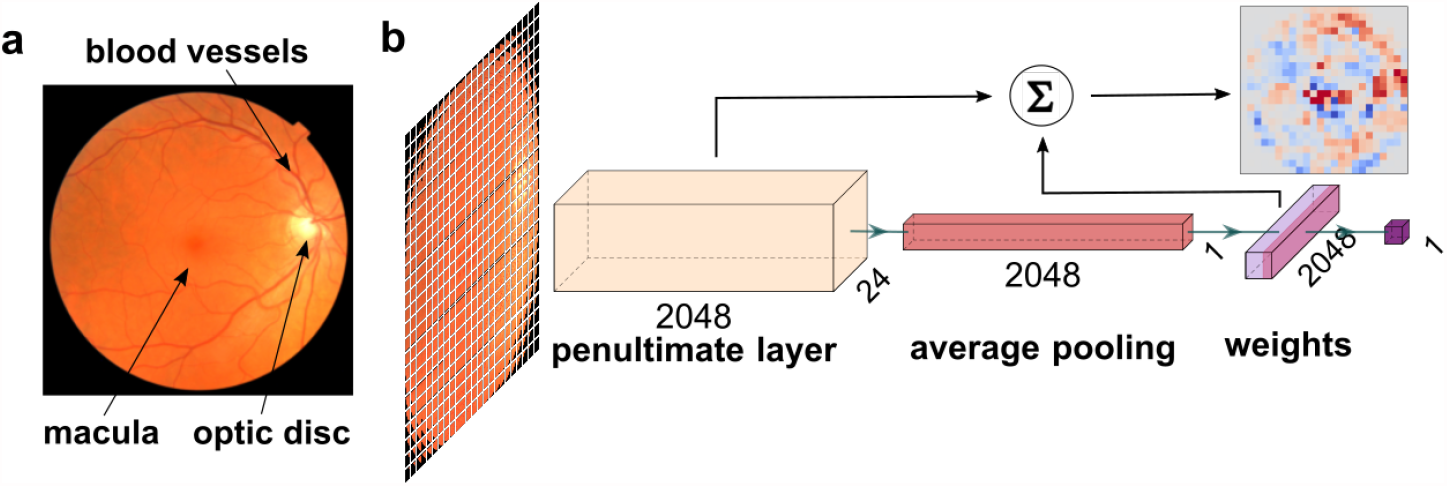
A sketch of the gender prediction via BagNet33. **(a)** Example fundus image from the UK Biobank. The optic disc is the bright spot on the right, the macula is the slightly darker spot in the middle and the blood vessels are extending from the optic disc in darker red. **(b)** The BagNet33 extracts 2048-dimensional feature vectors from 33 ×33 patches and stores them in the penultimate layer. Via spatial average pooling and a linear classifier, it then forms the final predictions for the gender. The same linear classifier can be applied directly to the feature representation in the penultimate layer to compute the local evidence, which can be visualized as a saliency map. Plotted with PlotNeuralNet [14].

## 2 Related Work

Previous work on gender prediction from fundus images have used either standard DNN architectures or simple logistic regression on top of expert-defined features. For example, [25] trained Inception-v3 networks on the UK Biobank dataset to predict cardiovascular risk factors from fundus images and found that DNNs were also capable of predicting the patient’s gender (AUC = 0.97). A similar network was used by [9]. In both studies, the authors computed posthoc saliency maps to study the features driving the network’s decisions. In a sample of 100 attention maps, [25] found that the optic disc, vessels, and other nonspecific parts of the images were frequently highlighted. However, this seems to be the case for almost all the dependent variables and it is very hard to derive testable hypotheses for gender specific differences. Likewise, [9] manually inspected a sample of occlusion maps and concluded that DNNs may use geometrical properties of the blood vessels at the optic disc for predicting gender. More recently, [5] demonstrated that DNNs can predict gender not only from retinal fundus images but also from OCT scans, where the foveal pit region seemed most informative based on gradient-based saliency maps. Taking a different approach, [29] used expert-defined image features in a simple logistic regression model. Although the performance of their model was worse (AUC = 0.78), they found various color-intensity-based metrics and the angle between certain retinal arteries to be significant predictors, but most effect sizes were small.

BagNets provide a compromise between linear classifiers operating on expert-defined features [29] and high-performing DNNs [25, 9, 5], which require complex post-hoc processing for interpretability [22, 2]. In BagNets, a saliency map is also straightforward to compute by design, and it has been shown to provide more information about the location of class evidence than auxiliary interpretability methods [3]. Such native evidence-based maps returned by BagNets are interpretable as is, while standard saliency maps require fine-tuning and post-processing for compelling visualizations [2]. Thanks to these benefits, BagNets have also been used in the context of histopathological microscopy [24].

## 3 Methods

### 3.1 Data and preprocessing

The UK Biobank [27] offers a large-scale and multi-modal repository of health-related data from the UK. From this, we obtained records of over 84, 000 subjects with 174, 465 fundus images from both eyes and multiple visits per participant. Male and female subjects constituted 46% and 54% of the data, respectively. As a substantial fraction of the images were not gradable due to image quality issues (artefacts, high contrast, or oversaturation), we used the EyeQual networks [6] to filter out poor images. 47, 939 images (47% male, 53% female) passed the quality check by the EyeQual ensemble. We partitioned them into the training, validation and test sets with 75%, 10% and 15% of subjects, respectively, making sure that all images from each subject were allocated to the same set.

Additionally, we obtained 29 fundus images from patients (11 male, 18 female, all older than 47 years) at the University Eye Hospital with permission of the Institutional Ethics Board. We used these additional images as an independent test set. For all images, we applied a circular mask to capture the 95% central area and to remove camera artifacts at the borders.

### 3.2 Network architecture and training

We used BagNets [3] (Fig. 1b) and standard Inception-v3 [28] network as implemented in Keras [4]. In a BagNet, neurons in the final layer have a receptive field restricted to *q×q* pixels, where we used *q∈ {* 9, 17, 33*}*. The convolutional stack in the network extracts a 2048-dimensional feature vector for each *q× q* image patch. Patches were implicitly defined, with a stride for convolutions of 8 pixels for *q* = 33. Therefore local features were extracted for each patch on a 24 × 24 grid (Fig. 1b). A linear classifier combined these 2048 features to obtain the local class evidence which was then averaged across all image patches (average pooling layer).

All networks had been pretrained on ImageNet [26] by their respective developers. For our binary classification problem, we replaced the 1000-way softmax output layer with a single logistic output neuron (Fig. 1b). We initially trained only the output layer using the fundus images for 10 epochs. This was followed by fine-tuning all layers for 100 epochs. We used stochastic gradient descent (SGD) with the learning rate set to 0.01 and the batch size to 16. We used data augmentation via random rotations and flips, width and height shifts, random brightness, and random zooming operations. We picked the best epoch from the [95, 100] range based on the validation performance. We evaluated the final performance on both the test set and the data from the University Eye Hospital.

### 3.3 Generation of saliency maps

To compute saliency maps, we applied the weights **w** in the final classification layer of BagNet33 to the feature vectors, e.g. **x**, in its penultimate layer (Fig. 1b), yielding the local evidence (logits) for each patch via **w** · **x** =∑ _*i*_ *w*_*i*_*x*_*i*_. We clipped the resulting values to [−75, 75] for visualization purposes. The resulting saliency maps were 24 × 24 (Fig. 2).

**Fig. 2:**
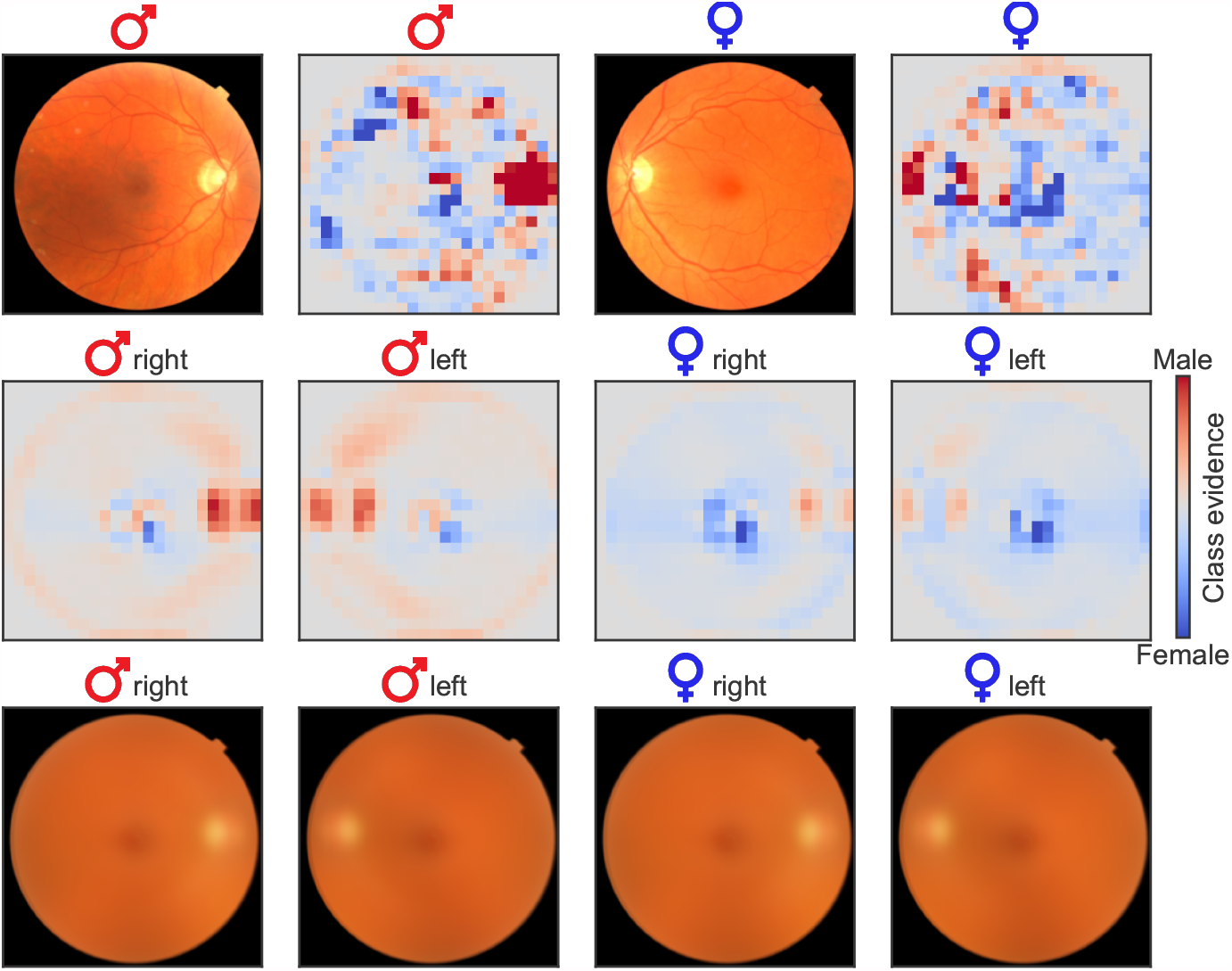
Saliency maps obtained by BagNet showing class evidence for each of the image patches on a 24 × 24 grid. Top row shows exemplary test images along with their saliency maps. Middle row shows the average saliency maps for correctly classified male and female patients. Bottom row shows the average fundus images corresponding to the middle row.

### 3.4 Embedding of image patches

To explore which image patches were informative for classification, we used t-Stochastic Neighborhood Embeddings (t-SNE) [20], a non-linear dimensionality reduction method. To embed the feature representations of *>*1, 000, 000 image patches extracted from the fundus images, we used FIt-SNE implementation [19] with uniform affinity kernel in the high-dimensional space across 15 nearest neighbours. We used PCA initialization to better preserve the global structure of the data and improve the reproducibility [16]. We used a heavy-tailed kernel *k*(*d*) = 1*/*(1 + *d*^2^*/α*)^*α*^ with *α* = 0.5 to emphasize cluster structure [17].

## 4 Results

We trained BagNets with three different receptive field sizes to predict patient’s gender from retinal fundus images based on the UK Biobank data. We evaluated their performances using prediction accuracy and the Area Under the Receiver Operating Characteristic curve (AUC) and compared to an Inception-v3 network (Table 1). BagNet33 and Inception-v3 performed on par with each other, while BagNet17 and BagNet9 performed worse. BagNet33 and Inception-v3 also generalized better to a new clinical dataset, albeit with a substantial drop in performance. Together, this suggests that the 33× 33 patches captured the relevant information for gender prediction better than smaller patches. Thus, for the remainder of the paper, we will focus our analysis on the BagNet33 (referring to it simply as BagNet).

**Table 1:** Gender prediction performances of DNNs

|  | TRAINING |  | VALIDATION |  | TEST |  | CLINICAL |  |
| --- | --- | --- | --- | --- | --- | --- | --- | --- |
|  | ACC. | AUC | ACC. | AUC | ACC. | AUC | ACC. | AUC |
| InceptionV3 | 92.99 | 0.98 | 83.87 | 0.92 | 82.97 | 0.91 | 62.07 | 0.78 |
| BagNet33 | 93.44 | 0.98 | 85.48 | 0.93 | 85.26 | 0.93 | 72.41 | 0.70 |
| BagNet17 | 86.66 | 0.94 | 82.30 | 0.90 | 82.11 | 0.90 | 37.93 | 0.51 |
| BagNet9 | 82.41 | 0.92 | 79.95 | 0.89 | 80.57 | 0.90 | 41.38 | 0.45 |

We inspected saliency maps for gender prediction computed by evaluating the classifier on each feature representation in the penultimate layer (Fig. 2, top). In a typical male example, we found that the optic disc provided high evidence for the male class, along with more scattered evidence around the major blood vessels. For a typical female example, high evidence was found for the female class in the macula. Averaging the saliency maps across all correctly classified male/female test images confirmed that the BagNet relied on the optic disc and the blood vessels to identify male images and on the macula to identify female ones (Fig. 2, middle).

Interestingly, the individual and the average saliency maps also showed that the optic disc patches tended to always provide male evidence, to some extent even in correctly classified female images. Similarly, the macula patches tended to provide female evidence, even in correctly classified male images. The BagNet could nevertheless achieve high classification performance after averaging the class evidence across all patches.

As a sanity check, we show the averaged fundus images across all correctly classified male/female images in the bottom row of Fig. 2. These average images are nearly identical across genders, demonstrating that it is not the location, the size, or the shape of the optic disc or macula that drive the BagNet predictions.

To further explore the structure of local image features informative about gender, we embedded the 2048-dimensional feature representation of each image patch into 2D using t-SNE and colored them by the provided class evidence (Fig. 3). We found that most image patches provided only weak evidence for either class, but some distinct clusters of patches had consistently high logits. We further explored these clusters and found that they consistently showed the optic disk with blood vessels (**a** and **c**) or the macula (**b** and **d**), in line with the saliency maps computed above (Fig. 2). However, even though the clusters **a** and **b** consistently provided evidence for the male class, patches in these clusters occurred in true female and male fundus images alike (67% and 62% patches from male images, respectively). Similarly, clusters **c** and **d** provided evidence for the female class but yet came from male and female fundus images (39% and 45% patches from male images, respectively).

**Fig. 3:**
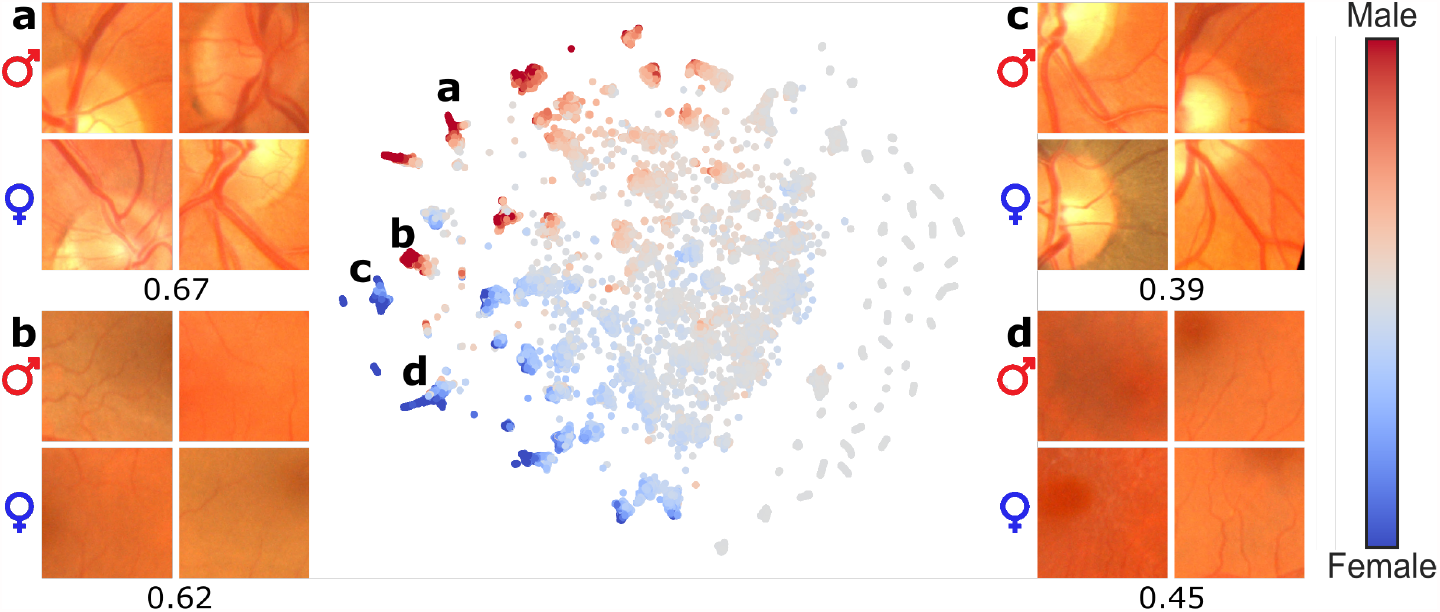
Visualization of image patches and associated class evidence via t-SNE. 213,696 patches extracted from 371 correctly classified test images (using training set images yielded a similar embedding; not shown). Four patches with high evidence (two male, two female) are shown from each of the four highlighted clusters. The fraction of male patches in each of these clusters is given below the corresponding exemplary patches. The colors show the logit class evidence. Note that the color does not indicate the correct label of each patch.

This raised the question of whether the BagNet’s decisions were mostly driven by (i) male/female images having individual patches with stronger male/female evidence; or (ii) male/female images having a larger number of patches with male/female evidence (Fig. 4). We found that both factors played a role in determining the final gender predictions, but the fraction of male/female patches seemed to be a stronger factor: Cohen’s *d* = 1.82 and *d* = 1.63 for the difference in fraction of male (logit value *>*50) and female (logit value *<−* 50) patches between genders, vs. *d* = 0.77 and *d* = 0.76 for the difference in the logit value of the most male and the most female patch. Thus, female images contained more patches providing strong female class evidence, and vice versa for male fundus images.

**Fig. 4:**
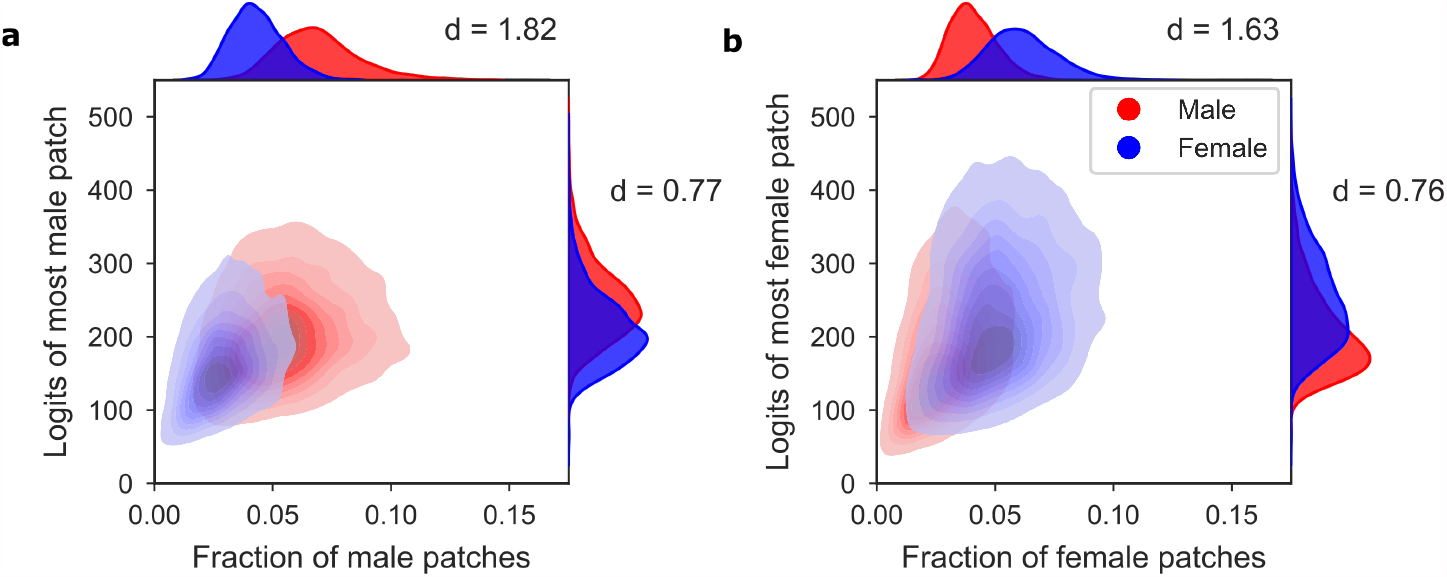
Two factors determine the gender predictions of the BagNet: the maximal strength of evidence and the frequency of strong evidence. **(a)** Kernel density estimate of all male (red) and female (blue) test set images. Horizontal axis: fraction of male patches, defined as having logit values above 50. Vertical axis: the absolute logit value of the most male patch. **(b)** The same for patches providing female evidence (logit values below −50).

## 5 Discussion

In summary, we argued that the BagNet architecture is particularly suitable for medical image analysis, thanks to its built-in interpretability. Here we used BagNets to investigate the high accuracy of DNNs in gender prediction from retinal fundus images. BagNet33 achieved a performance similar to Inception-v3 despite having a much simpler architecture and using only local image features for prediction. This suggested that local features are sufficient for gender prediction and the global arrangement of these features is not essential for this task.

In BagNets, saliency maps can be readily computed without auxiliary gradient-based methods or layer-wise relevance propagation [22]. We used the native saliency maps of BagNets and a two-dimensional t-SNE embedding of image patches to identify the most informative regions for the gender prediction task in fundus images. This allowed us to go beyond the previous reports [25, 9] and for the first time to provide conclusive evidence that the optic disk region contains features used to inform a male prediction and the macula region for a female prediction. We found that both the frequency of informative male/female patches and — albeit to a lesser degree — the strength of the most informative male/female patches were important factors for gender prediction by BagNets.

It is, however, not the case that the optic disc in males is substantially larger than in females, as can be seen in the average fundus images shown in Fig. 2. The relative optic disc and macula sizes, shapes, brightness levels, etc. seem all to be roughly the same for both genders. Instead, our results suggest *structural but localized* differences in the male and female retinas, mainly within the optic disc and macula regions. This is supported by the previous findings showing that the retinal nerve fibre layer in the optic disk is slightly thicker in females [18] and that the macula is slightly thinner [5] and wider [8] in females. However, these previously reported gender differences have small to moderate effect sizes (Cohen’s *d* = 0.11, *d* = 0.52, and *d* = 0.17 respectively for the comparisons referenced above; computed here based on reported means and standard deviations) and it is unclear if they alone can explain the BagNet performance.

Therefore, future work is needed to understand what exactly it is that allows the network to assign high male evidence to the optic disc patches from male patients and high female evidence to the optic disc patches from female patients. In this sense, the results presented here do not provide the final solution to the gender prediction mystery. Nevertheless, we believe that our results make a step in the right direction as they demonstrate structural but localized gender differences and reduce the problem complexity down to specific small patches of the fundus image that can be further analyzed separately.

We believe that BagNets may also be more widely applicable for clinically relevant diagnostic tasks involving medical images in ophthalmology and beyond, provided that they are coupled with reliable uncertainty estimation [1]. In many cases, pathologies often manifest in localized regions, which can be readily picked up by BagNets. For example, BagNets could be used to further explore clinically relevant changes underlying progressive diseases such as diabetic retinopathy. The interpretable architecture of BagNets may increase the trust of clinicians and patients, which is a critical issue for adoption of deep learning algorithms in medical practice [11].

## Data Availability

The main collection of retinal fundus images were obtained from the UK Biobank. Due to the Material Transfer Agreement of UK Biobank, we can not share the data, but researchers can apply for access and subsidized fees are available.

## Acknowledgements

We thank Wieland Brendel for his support with BagNets. This research was supported by the German Ministry of Science and Education (BMBF, 01GQ1601 and 01IS18039A) and the German Science Foundation (BE5601/4-2 and EXC 2064, project number 390727645). Additional funding was provided by Novartis AG through a research grant. The funding bodies did not have any influence in the study planning and design.

